# Gastroenterology Procedures Generate Aerosols: an Air Quality Turnover Solution to Mitigate the Risk

**DOI:** 10.1101/2020.08.21.20178251

**Authors:** Marc Garbey, Guillaume Joerger, Shannon Furr

## Abstract

The growing fear of virus transmission during the 2019 coronavirus disease (COVID-19) pandemic has called for many scientists to look into the various vehicle of infection, including the potential to travel through aerosols. Few have looked into the issue that gastrointestinal (GI) procedures may produce an abundance of aerosols. The current process of risk management for clinics is to follow a clinic-specific HVAC formula, which is typically calculated once-a-year and assume perfect mixing of the air within the space, to determine how many minutes each procedural room refreshes 99% of its air between procedures when doors are closed. This formula is not designed to fit the complex dynamic of small airborne particle transport and deposition that can potentially carry the virus in clinical conditions. It results in reduced procedure throughput as well as an excess of idle time in clinics that process a large number of short procedures such as outpatient GI centers.

We present and tested a new cyber-physical system that continuously monitors airborne particle counts in procedural rooms and also at the same time it automatically monitors the procedural rooms’ state and flexible endoscope status without interfering with the clinic’s workflow. We use our data gathered from over 1500 GI cases in one clinical suite to understand the correlation between air quality and standard procedure types as well as identify the risks involved with any HVAC system in a clinical suite environment. Thanks to this system, we demonstrate that standard GI procedures generate large quantities of aerosols, which can potentially promote viral airborne transmission among patients and healthcare staff. We provide a solution for the clinic to improve procedure turnover times and throughput, as well as to mitigate the risk of airborne transmission of the virus.

## 1 Introduction and Motivation

During a pandemic, such as the one of COVID-19 [1], a careful management of elective procedures must ensure that patients and staff do not take on excessive risk [2]. At the same time it is critical to maintain the clinic’s capacity in order to keep the population healthy with preventive medicine and therapy [3]. This is particularly true for GI centers [4, 5] who are working restlessly to prevent a large number of patients from developing cancer and deliver therapy as early as needed.

We focus our paper on routine GI procedures typically practiced in high volume outpatient centers, but lessons learned from that context may serve other types of clinical workflow. One can observe multiple factors impacting workflow due to COVID-19:

- Turnover time: GI procedures generate significant aerosol [34], and virus tests for patients may have limited reliability [26] [17]; consequently, some facilities implement longer turnover than usual to make sure that the air has been completely recycled in the procedural room.
- Volume of patients is impacted by multiple factors related to COVID-19:

– Patients do not feel safe coming to the hospital and may delay their procedure or cancel more often – in particular, when it is a routine exam. Clinics have almost no-way to explain how they manage COVID-19 to their patients.
– Patients are required to cancel their procedure if their COVID-19 test is positive. This means that there will be more patient coming to clinics once the pandemic has slowed down.
– Depression and anxiety induced by the pandemic may increase the number of patients who require GI procedures.
- Staff working conditions are impacted by COVID-19:

– First, the number of staff available including nurses and techs may decrease significantly, either because of sick leaves, children’ responsibilities, or redeployment of staff to manage the pandemic, last but not least staff can also be stressed by the situation if no action is taken from the management.
– Second, some of the routine tasks may take longer because of the additional personal protective equipment (PPE) requirements and new protocols for safety. This should be true for a scope reprocessing facility, which has a very humid environment and has very high airborne particle counts, as measured in our data set.
- Maintenance and external services: if any equipment breaks, maintenance may take much longer during a pandemic.

We have specifically developed mathematical and cyber-physical tools to address some of the workflow issues via simulation calibrated on unbiased, real-time, autonomous measurement of clinic performances [10–13,16,19].

Air quality in hospital and clinic environment should be revisited in the light of new risk with the pandemic. The mechanism of airborne transmission of the severe acute respiratory syndrome coronavirus 2 (SARS-CoV-2) in closed buildings [8,23] is still poorly understood. Surgical smoke can act as a carrier for tissue particles, viruses, and bacteria [32] and has been studied for some time [6,14,15,24,25,29,30]. It was recently shown in a study that air quality, especially concentration of fine particles, is associated with an increase in COVID-19 mortality [36]. Respiratory protection devices are used to protect staff in healthcare facilities with various degrees of success [7, 21]. We proposed recently a system approach to study the multiple scales of hospital air quality in relation to human factor and described the method in [12].

We focus our study specifically on two new aspects of the workflow’s safety and efficiency. First, we show quantitatively that standard GI procedures, such as colonoscopy and esophagogastroduodenoscopy (EGD), generate a significant number of aerosols. Then, we propose a practical solution to improve the turnover process from both the perspective of safety and efficiency. Risk mitigation techniques balance the benefit of the medical procedures with risk of contamination. The risk of contamination will be lower provided that the best practice and new guidance directive of the CDC [2] [34] are scrupulously followed.

We advocate that it is useful to get real-time indication of risk level during the clinical day to adapt behavior accordingly in a similar way that staff protect them self of the effect of radiation with X-ray imaging. We attempt to demonstrate a technique towards that goal in a GI clinic environment.

The paper is organized as follows: Section 2 describes our method to construct the infrastructure and assess particle count level in real-time to guide the turnover process between procedures; Section 3 gives the main results and solution to our initial goal in supporting management; Section 4 discusses the benefit and limitation of our method and concludes with further potential development.

## 2 Methods

We start with some key observations that were instrumental in developing the method, then go into the infrastructure of our method, and end with the measurements supporting our method.

### 2.1 Key Observations

We begin with a practical example of what we have been able to measure in a clinical environment during the COVID-19 pandemic. We illustrate the correlation between procedure and room state along with airborne small particle counting in the procedural room. Figure 1 shows the procedure state (top graph) and particle counts (bottom graph) in a procedural room during a clinical day in an outpatient center. For simplicity, we chose to start with a light activity day in the procedural room with only three patient procedures. The top figure gives the room state: the red step function is 1 during a colonoscopy, 2 during an EGD. The first patient went through an EGD from 7:40 am to 8:00 am, the second patient had a colonoscopy starting around 9:00 am, and the third patient had a colonoscopy around 11:00 am. We define the turnover time to be the time between one patient leaves the procedural room and the next patient enters. One will notice that the turnover time is very large, i.e. about 1 hour for the first interval between the first and second patient procedures, and 1.5 hours for the second interval between the second and third patient procedures. This is very unusual since turnover time in this clinic prior to COVID-19, was about 5 to 10 minutes on average. The 3 procedures’ duration are however similar to procedure durations before COVID-19, i.e. about 10 minutes for an EGD and 20 minutes for a colonoscopy. The black step function in the top part of Figure 2 shows the state of the endoscope that can help us monitor the state of the room. Initially, the step functions stay at zero value until 7:00 am, which corresponds to the endoscopic tower not in use or powered up. When the step function is 1, the flexible endoscope is not plugged to the endoscope tower, but the endoscope tower has been powered on. When its value is 2, it means that the flexible endoscope has been plugged in but is not in use for the procedure. When its value is 3, the flexible endoscope is in use for the procedure diagnostics and/or therapy. The scope state follows the cycle [1 2 3, 1 2 3…] until the last procedure is finished. The clinical day usually ends around 4:00 pm and the endoscope tower is eventually powered down, so the scope state will return to 0. One may notice that the flexible endoscope was ready to be used long before the patient procedure started for patient 1 and 3, but not for patient 2.

**Figure 1.**
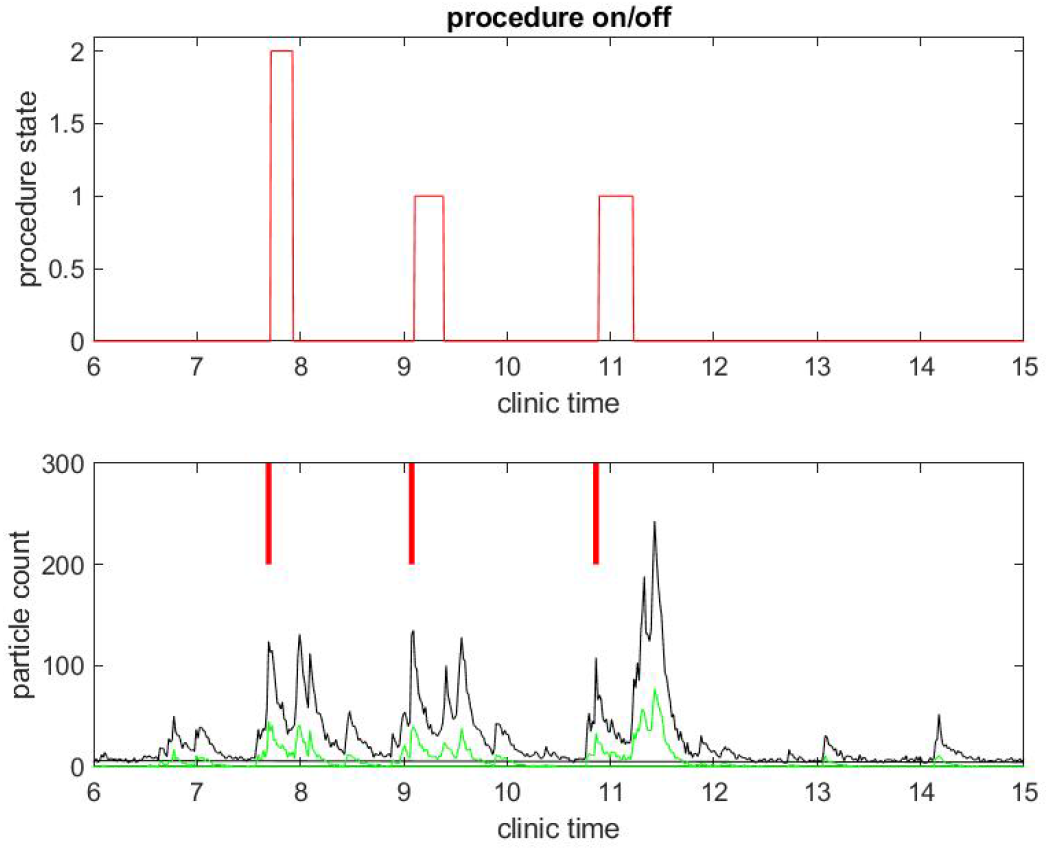
Example of particle counts during a clinical day in a procedural room. The upper graph displays the procedure state, 1 for colonoscopy and 2 for EGD procedure. The lower graph displays small particle counts (black) and large particle counts (green), along with red bars for marking when the procedure from the upper graph began.

**Figure 2.**
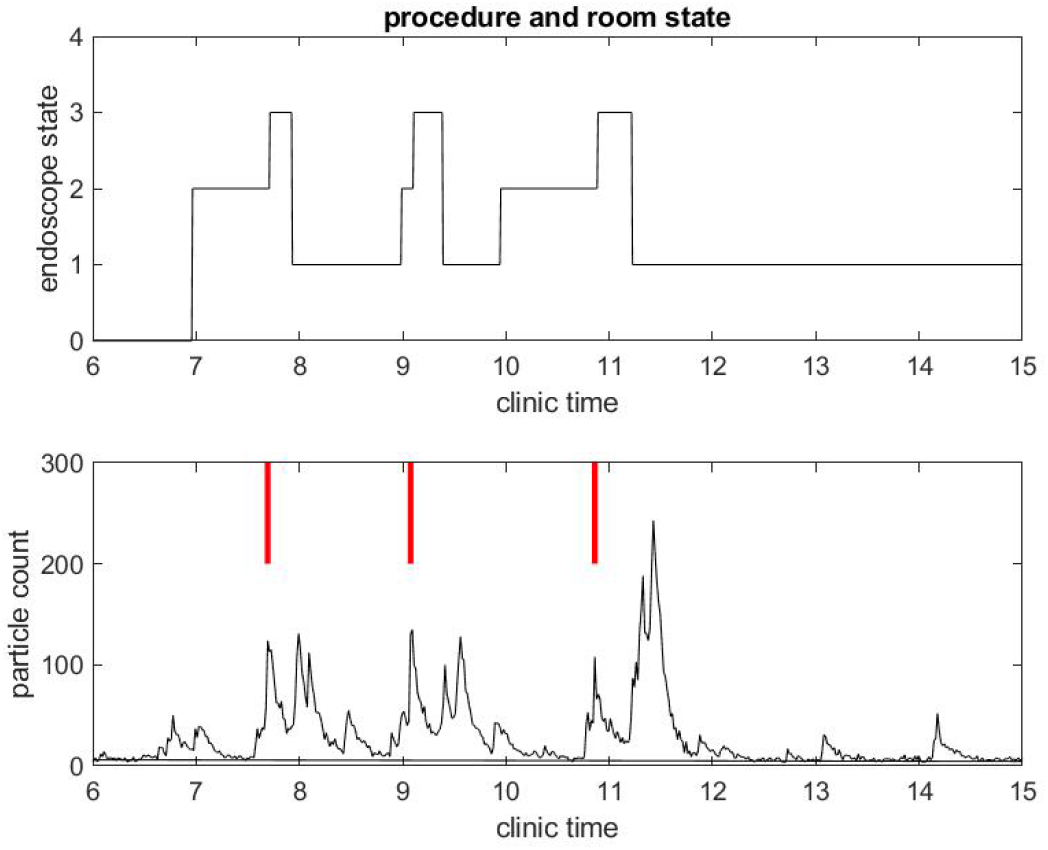
Example of small particle count during a clinical day in a procedural room. The upper graph displays the endoscope state, 0 for endoscope tower not in use, 1 for endoscope tower powered on, 2 for endoscope plugged in but not in use during procedure preparation, 3 for in use during procedure. The lower graph displays the small particle count only, with red bars marking when the procedure from the upper graph began.

We give context on the procedural activity to better explain the bottom graph that displays a real-time measurement of particle count with an off-the-shelf device Dylos DC 1700, Dylos Corp (Riverside, CA, USA) which we have already used extensively in our air quality study of surgical suites [12]. Following the work of Setti et al., we assume that airborne diffusion of infected droplets from person to person can occur at a distance greater than two meters [28]. In the unfortunate case a GI procedure is practiced on an asymptomatic COVID-19 patient who was not tested positive, contamination by airborne particles generated by the procedure itself might be an issue. The staff present during the procedure need to wear appropriate PPE. The next patient might be exposed as well. While the patient stays on its own bed and the room is cleaned in between procedure, if the air contamination has not been eliminated by the Heating, Ventilation and Air Conditioning (HVAC) system [18] a contamination could still be possible.

The Dylos laser particle counter gives an average particle count every minute in a unit system with units *u_d_* that correspond to particles per 0.01 cubic foot, or particles per 0.00028 cubic meter (1 cubic foot = 0.0283168 cubic meter). According to smartAir (http://smartairfilters.com/cn/en/), the Dylos system’s output is highly correlated (r = 0.8) to a “ground true” measurement provided by a high-end system, such as the Sibata LD 6S that is claimed to be accurate within 10% in controlled laboratory conditions. According to smartAir, the Dylos system seems particularly accurate at the lower concentration ends, which is of interest for this study’s purpose. Semple *et al*. [27] also compared the Dylos system with a more expensive system: the Sidepak AM510 Personal Aerosol Monitors (TSI, Minnesota, USA). They concluded that the Dylos system’s output agrees closely with the one produced by the Sidepak instrument with a mean difference of 0.09 *µg/m*^3^.

The black curves in the bottom graphs of Figure 1 and Figure 2 track particles of small size in the range from 0.5 to 2.5 *microns*, which are the sizes of most biological materials. The green curves in Figure 1 track particles of larger size (greater than 2.5 *microns*). We focus on small particles since they stay airborne longer than large particles and may be the main source of potential contamination. A Small Particle Count (SPC) below 50 *u_d_* is considered to be *excellentquality* according to the Dylos manufacturer. As the particle count number goes up, air quality degrades. In all of our measurements, we observed that the SPC is less than 50 *u_d_* when the procedural room is empty with the door closed: this shows the efficiency of the HVAC system that evacuates particles as it is expected. We will consider this level of particle count as the baseline.

A few observations can be taken from Figure 1. As mentioned above the SPC is (far) below 50 *u_d_* until 6:30 am, which shows indirectly that the door of the procedural room was closed until then. As soon as people come in and out of the room or the door stays open, the particle counts go up. For example, the flexible endoscope was plugged in the procedural room around 7:00 am according to Figure 2, and so we observe a small peak in the SPC around that time. To visually reference on the particle count graphic when the procedure started, we add a vertical red segment on Figure 1 and Figure 2: we can then observe clearly how air quality changes quickly with the start of the procedure. We see systematically a peak in the particle count at about the same time the procedure starts, i.e. when the flexible endoscope is introduced inside the patient; the particle count right before that time is much lower, while both the patient and the team have been in the room for several minutes and the door of the procedural room has been closed. So this peak is not relative to people already inside the room but rather the procedure itself. This observation confirms on the example that both EGD and colonoscopy are generating aerosol significantly, as listed in the review of Tran *et al*. [31].

The same observations can be done in a different procedural room the same day that had six procedures occur instead of three, as shown in Figure 3 and Figure 4. Let us notice that this was for five patients, since the fourth patient got an EGD immediately followed by a colonoscopy. Once again regarding the second example, the duration of the procedures were about standard, but the turnover time between procedures was fairly large compared to pre-COVID workflow statistics.

**Figure 3.**
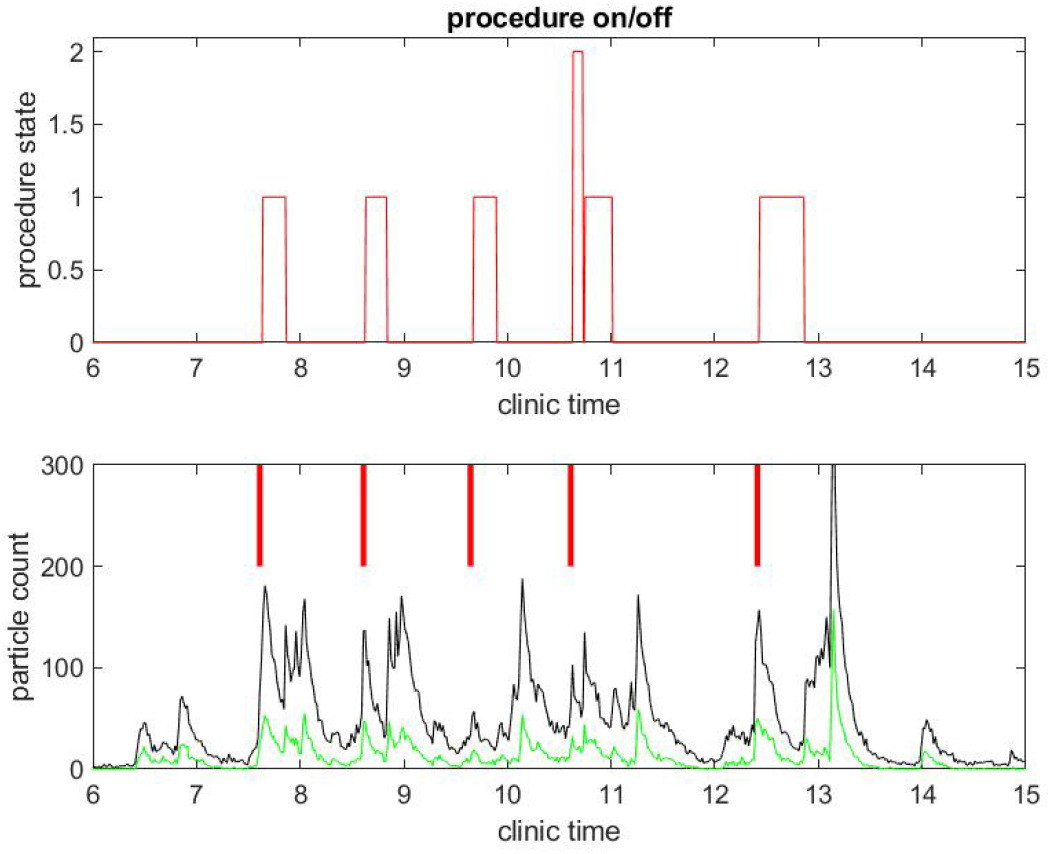
Example of particle counts during a clinical day in a procedural room. The upper graph displays the procedure state, 1 for colonoscopy and 2 for EGD procedure. The lower graph displays small particle counts (black) and large particle counts (green), along with red bars for marking when the procedure from the upper graph began.

**Figure 4.**
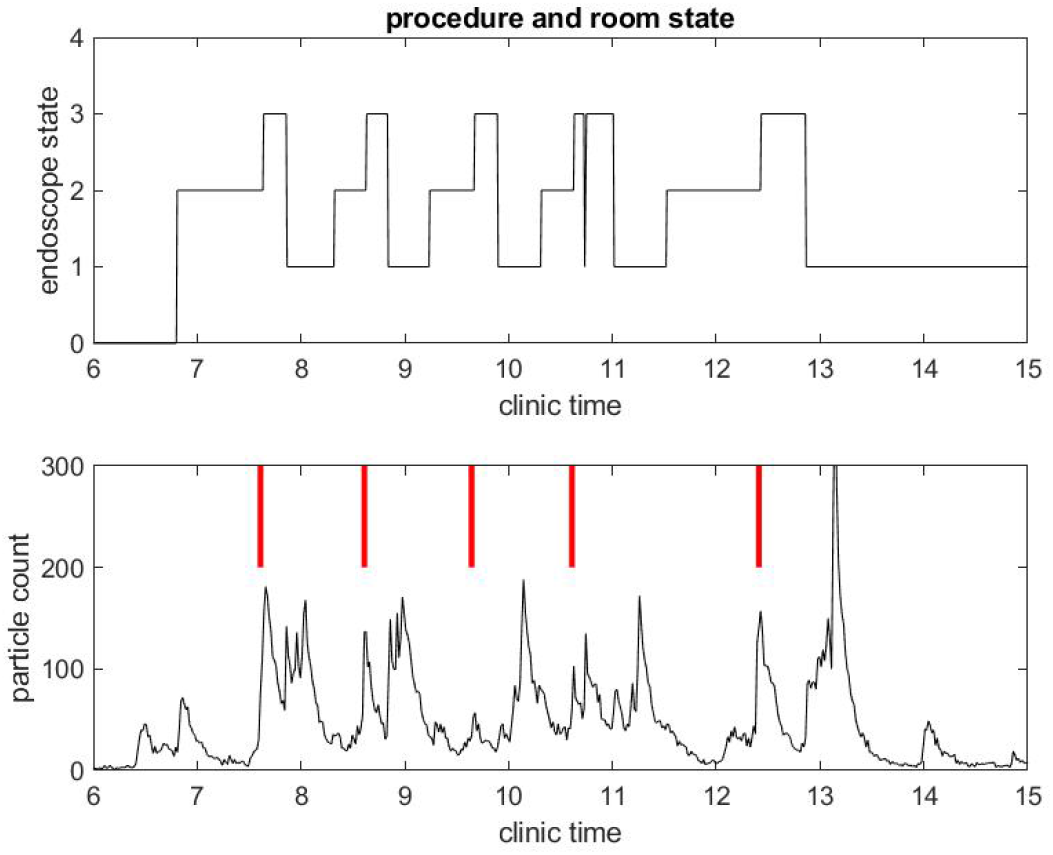
Example of small particle count during a clinical day in a procedural room. The upper graph displays the endoscope state, 0 for endoscope tower not in use, 1 for endoscope tower powered on, 2 for endoscope plugged in but not in use during procedure preparation, 3 for in use during procedure. The lower graph displays the small particle count only, with red bars marking when the procedure from the upper graph began.

Another remarkable feature of the SPC measure is that it catches the time at which the janitorial team arrives into the procedural room to clean surfaces: this process generates a lot of aerosol with the evaporation of disinfectant product. It can even be seen that at the end of the day, the cleaning is done more extensively in both procedural rooms and generates the largest peak of particle counts.

We now show a systematic way of analyzing these results using the insight we gained from these observations. To provide a systematic measurement of air quality versus procedures, we first build an infrastructure.

### 2.2 Infrastructure

We have equipped each procedural room of a high-volume GI outpatient center with two sensors:

1. A ”computer vision sensor” that checks the endoscope’s video view and automatically retrieves the state of the endoscope in real-time, as shown in the top graphs of Figure 1 and Figure 2. It is relatively easy to obtain these states most of the time. However, we did extensive development on the algorithm to get a robust and accurate measure that holds an accuracy of timing of one minute.
2. A Dylos air quality sensor attached to the wall of the procedural room at about 1.5 m above the floor: these procedural rooms are relatively small compared to surgery operating rooms. We found in our previous CFD calculation and measurement with multiple air sensors [12], that air mixing takes less than a minute to give a relatively uniform SPC in standard surgical operating room. Due to the smaller size of the GI procedural room and the efficient HVAC system present in them, particle count should have little time lag compared to the source of particles (the staff or the patient). Further, we found the location of the sensor not very sensitive unless it is set at some corner of the room behind equipment that obstructs the airflow through the sensor.

Both sensors provide digital information in close-to-real-time which is then sent over the wireless local area network (WLAN) of the clinic to a server that archives the time series into a database. The server is behind the firewall of the clinic’s WLAN, so all data and network communications are protected. The data retrieved by our system contains no patient information: the flexible endoscopy video feed does not leave the endoscope tower nor is saved onto our system at any point. A significant amount of engineering development is needed in order to have a stable system that maintains an uninterrupted connection with a very low probability of information loss.

### 2.3 Measurement

We have collected the time series, demonstrated in the two examples displayed in Figure 1 to 4, over a period of about 50 consecutive clinical days in 7 procedural rooms simultaneously. Overall, we collected a data set with 1213 colonoscopies and 594 EGD.

To begin, we analyze the aerosol generated by the procedures with our SPC measurement. For those 301 patients who had a hybrid procedure (i.e. an EGD immediately followed by a colonoscopy) in our data set, the interval of time between the EGD and colonoscopy that follows is so short that the SPC during the colonoscopy might be impacted by the EGD. So, we removed the colonoscopies from these hybrid procedures in our study. For the 912 colonoscopies left and 594 EGD, we compute the average number of particles at different steps:

- Step 1: the 5 minute window prior to the procedure, when the medical team is already in the room and the door closed, with the air quality average during the whole procedure, noted *D_pre_*,
- Step 2: the first 5 minutes of the colonoscopy, and respectively the first 3 minutes of the EGD procedure, noted *D_start_*,
- Step 3: the rest of the procedure, noted *D_proc_*.

If the procedure generates aerosol, we expect that the average during Step 2 will be larger than during Step 1. Using our previous observation, our hypothesis is that in the first few minutes of the procedure (Step 2) the patient reacts to a foreign body inserted in his/her gastric path with more vigor than during the rest of the procedure. To verify the hypothesis, we also computed the average SPC during the procedure after the initial phase mentioned above (Step 3). The result section gives some statistics on these numbers and quantitatively shows that both colonoscopy and EGD procedures generate significant aerosols.

Next, we analyze the air quality of the procedural room during the turnover time between two patients. In order to avoid any interference and minimize the noise in the data set, we impose a number of criteria to restrict our analysis to a subset of ”procedure turnover” SPC signals. We assume:

1. that the procedural room goes through cleaning during a small interval of a few minutes right after the procedure is done,
2. then the door is closed and staff won’t open the door of the procedural room until the air quality is *excellent*, i.e. SPC has been below 50 *u_d_* for 5 minutes.

We use signal analysis on the SPC curves to keep only the signal from the Dylos sensor that would correspond to this hypothesis. Firstly, we keep only the data set with a sharp peak of particle counts that occurs not far after the procedure is done. Secondly, as soon as the door of the procedural room is closed, we observe that the particle count should decrease in the first approximation exponentially: We expect this exponential decay from the standard diffusion convection process in a closed space, with inlet/outlet flow corresponding to the HVAC system, that should be a dominant factor. Thirdly, we restrict ourselves to an interval of time that leads to a decay of particle count below 50 *u_d_*. Using all three criteria, we found a data set of 199 turnover air quality signals that we could analyze. The benefit of this approach is that we can properly compute the exponential decay of particle counts at turnover to check a few additional hypothesis without external interference; for example, we can show that large particle count decays faster than small particle count as expected. We can also compare how fast the HVAC system cleans out the air for each procedural room, and demonstrate that the formula presented by the CDC in [35] has strong limitations.

However, we will also report on a practical measurement of ”air quality turnover time” on a much larger data set to show that our result holds in more general circumstances. This is the most practical information for the manager of the clinic.

## 3 Results

Let us report first on our indirect measurement of aerosol generation for colonoscopy and EGD procedures, and then discuss the turnover times based on air quality levels.

### 3.1 Colonoscopy and EGD Generate Aerosols

Figure 5 shows the ratio of the mean SPC during the first 5 minutes of a colonoscopy divided by the mean SPC during the 5 minutes prior to the colonoscopy procedure 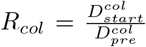. Usually during this time window (Step 1), the patient is already inside the procedural room along with the medical team, composed of the gastroenterologist, a registered nurse, anesthesiologist, and a technician who helps with the endoscope. We use this time window as a baseline to rate the particle count generated from the procedure itself, since the door is closed and the number of people who are in the procedural room typically stays the same. We observe that in the vast majority of the cases this ratio is above the *y* = *x* line. Figure 7 gives the histogram of the ratio *R_col_*, and shows a biased distribution with 2 as its maximum. According to our interpretation, these figures show that colonoscopy procedures generate a significant level of aerosol.

**Figure 5.**
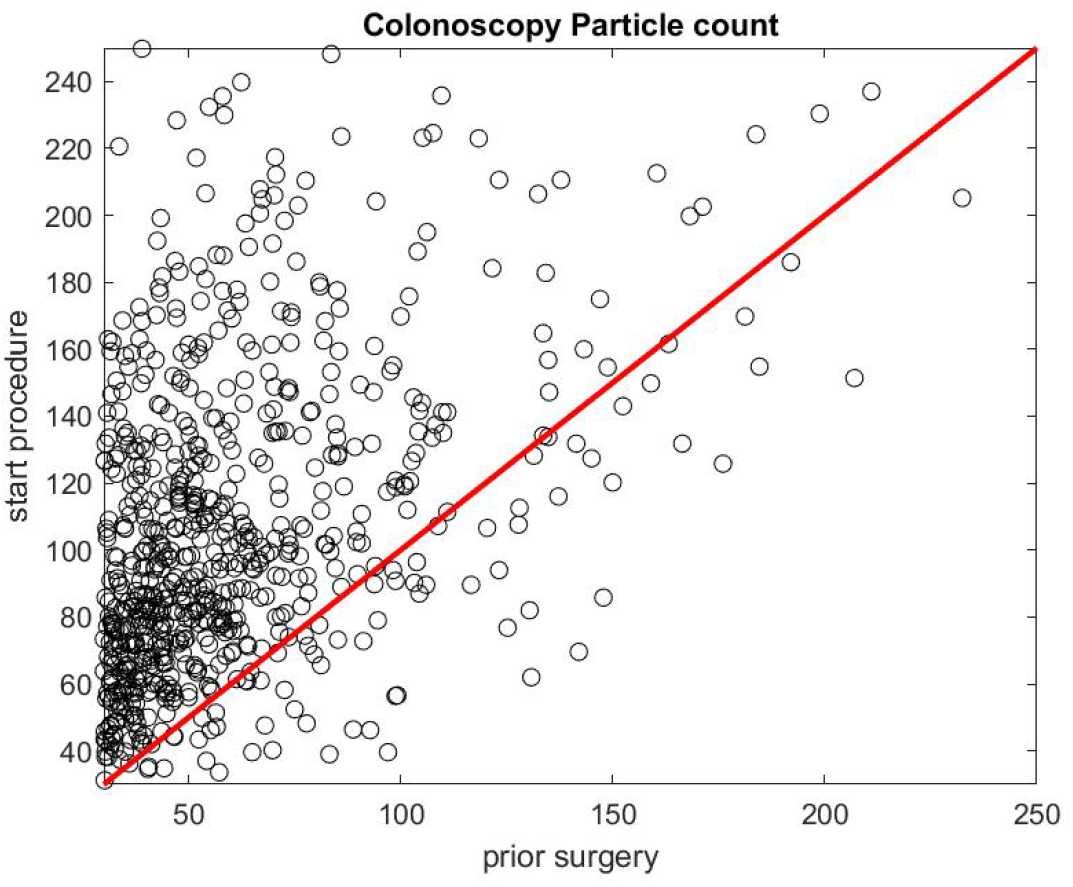
Particle count during the first 5 minutes of a colonoscopy *D_start_* versus particle count during the 5 minutes prior to the procedure *D_pre_*.

Figure 6 and Figure 8 report on similar results with EGD procedures with 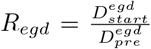. Because an EGD takes about half the time of a colonoscopy to complete, we look at the ratio of the mean SPC during the first 3 minutes only, 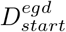, versus the mean SPC during the 5 minutes prior to the EGD procedure, 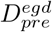. An EGD generates less particles in proportion to a colonoscopy during the initial phase of the procedure when accessing the anatomic region of interest with the endoscope. Figure 9 gives the distribution of the ratio *P_col_* of the mean SPC during the rest of the colonoscopy procedure (Step 3), and respectively Figure 10 for the EGD procedure *P_egd_*, versus the mean SPC prior to the procedure. The mean of that ratio over all colonoscopy procedures is 1.08, respectively 0.98 for EGD procedures. Of these procedures, 40% of them generate small particles during the procedure past the initial phase.

**Figure 6.**
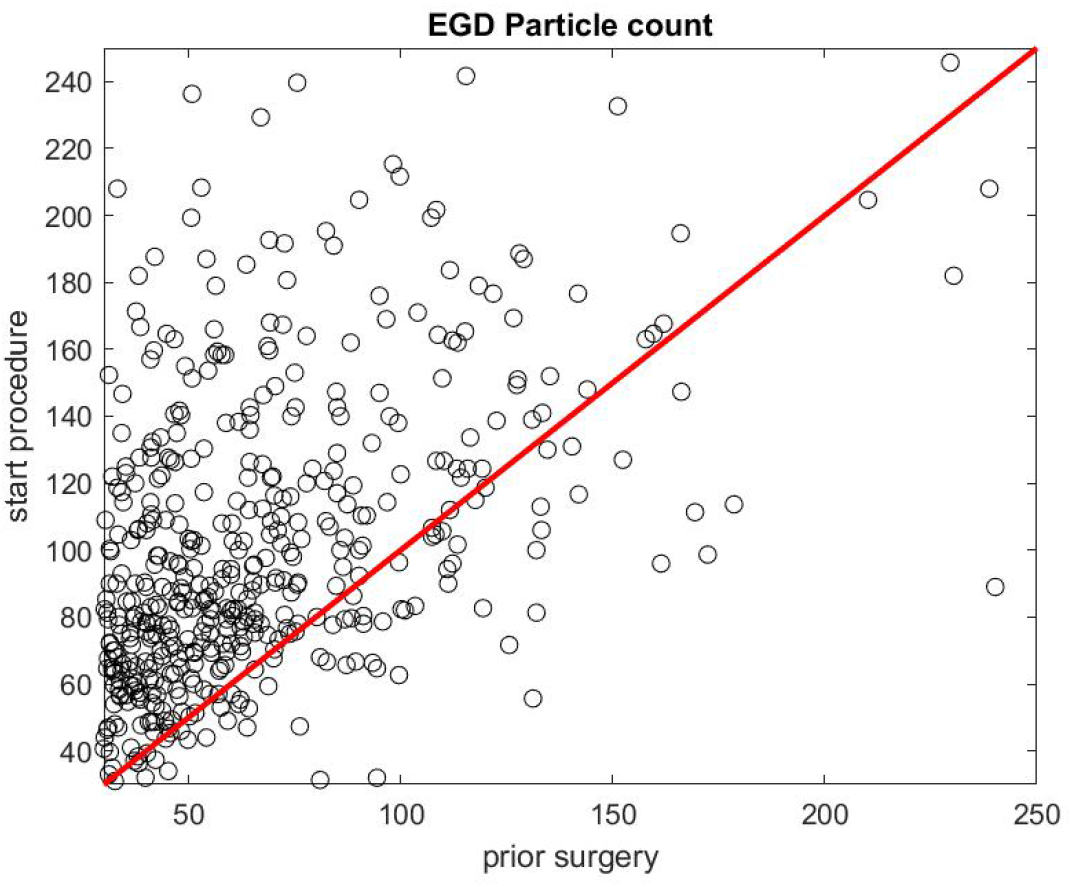
Particle count during the first 3 minutes of an EGD 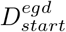 versus the particle count in the 5 minutes prior to the procedure 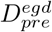.

**Figure 7.**
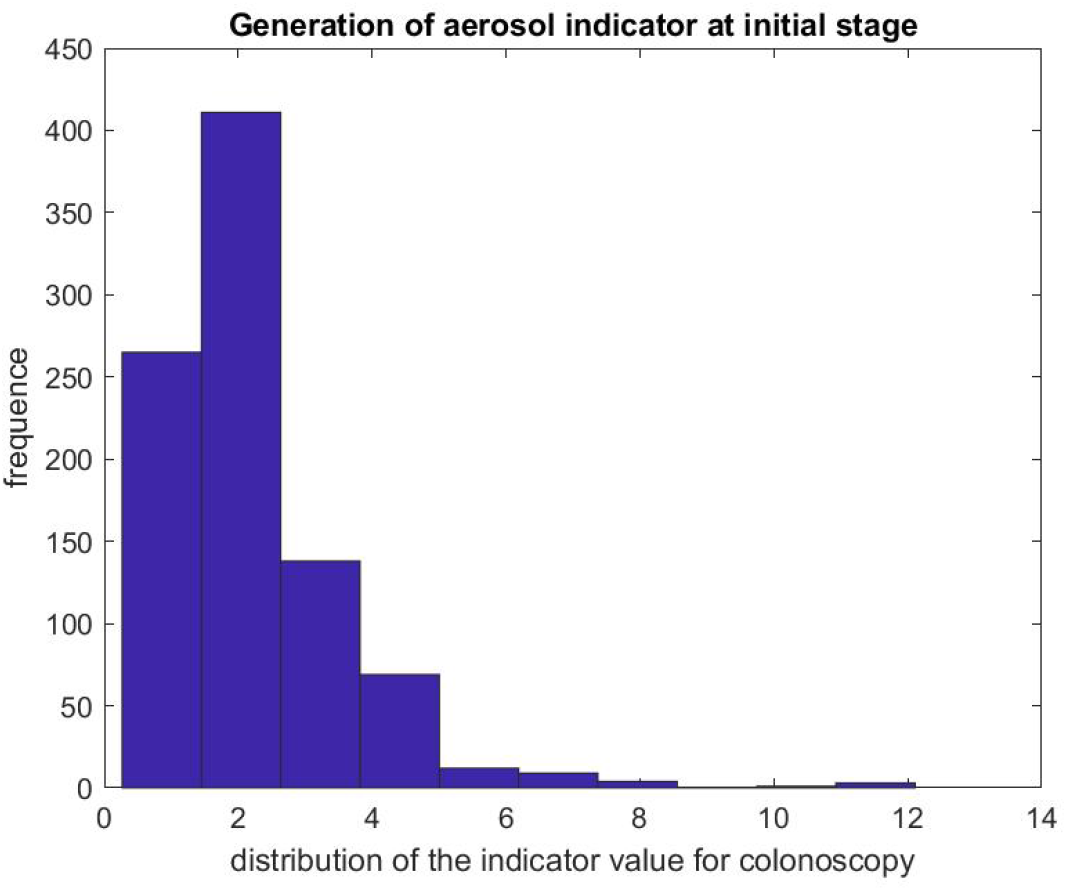
Indicator of aerosol generation for a colonoscopy during the first 5 minutes of the procedure. The figure represent the distribution of 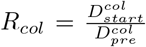

**Figure 8.**
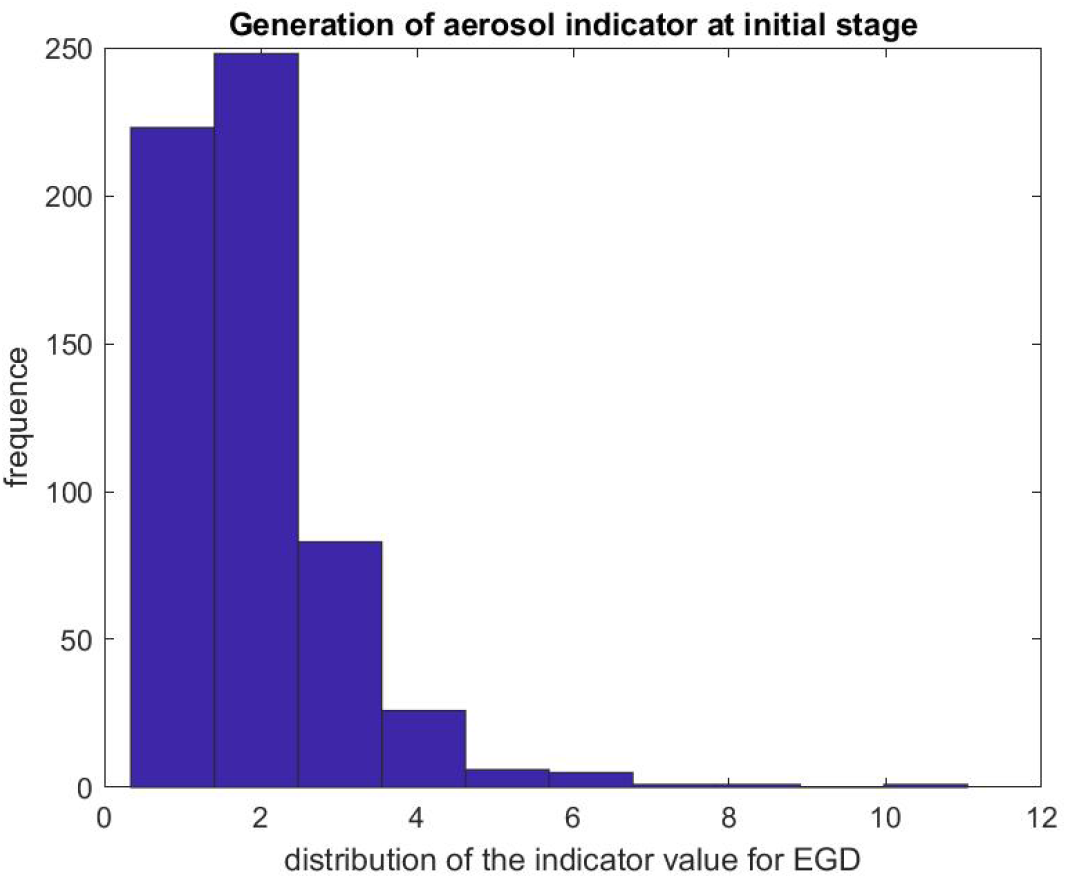
Indicator of aerosol generation for an EGD during the first 3 minutes of the procedure. The figure represent the distribution of the ratio 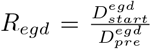

**Figure 9.**
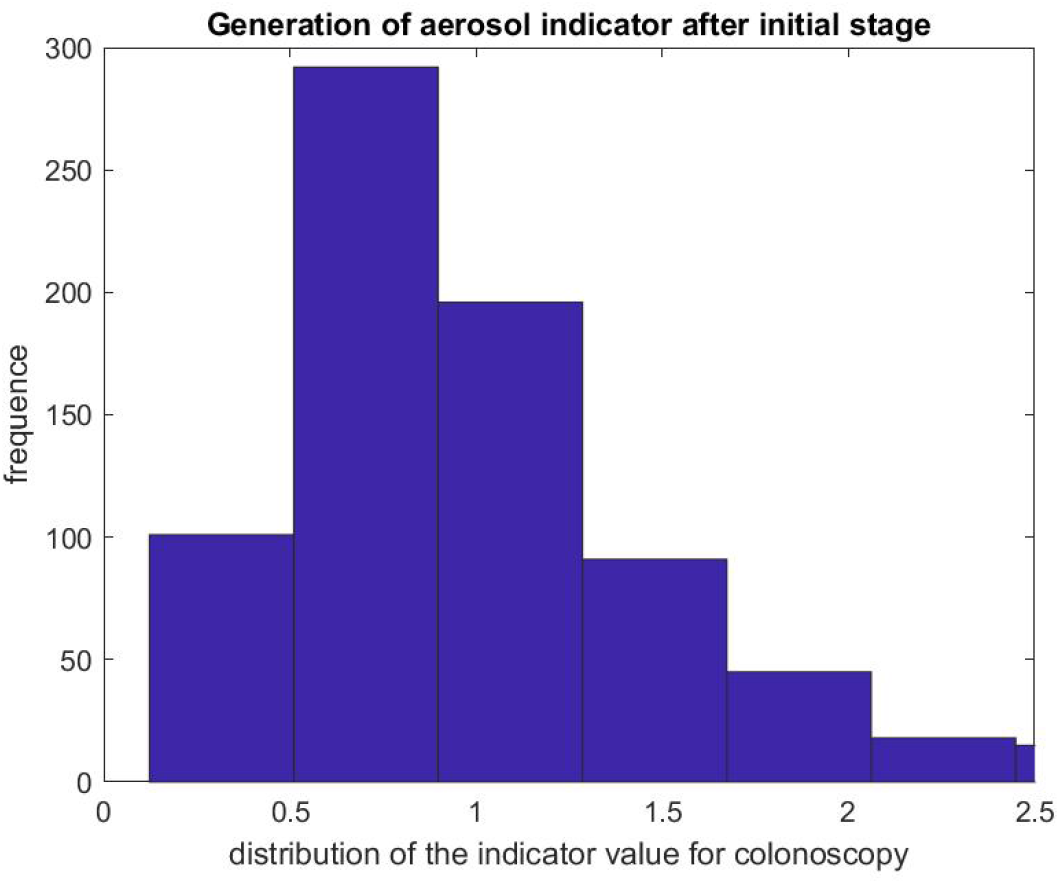
Indicator of aerosol generation for a colonoscopy after the initial phase. The figure represent the distribution of the ratio 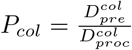

**Figure 10.**
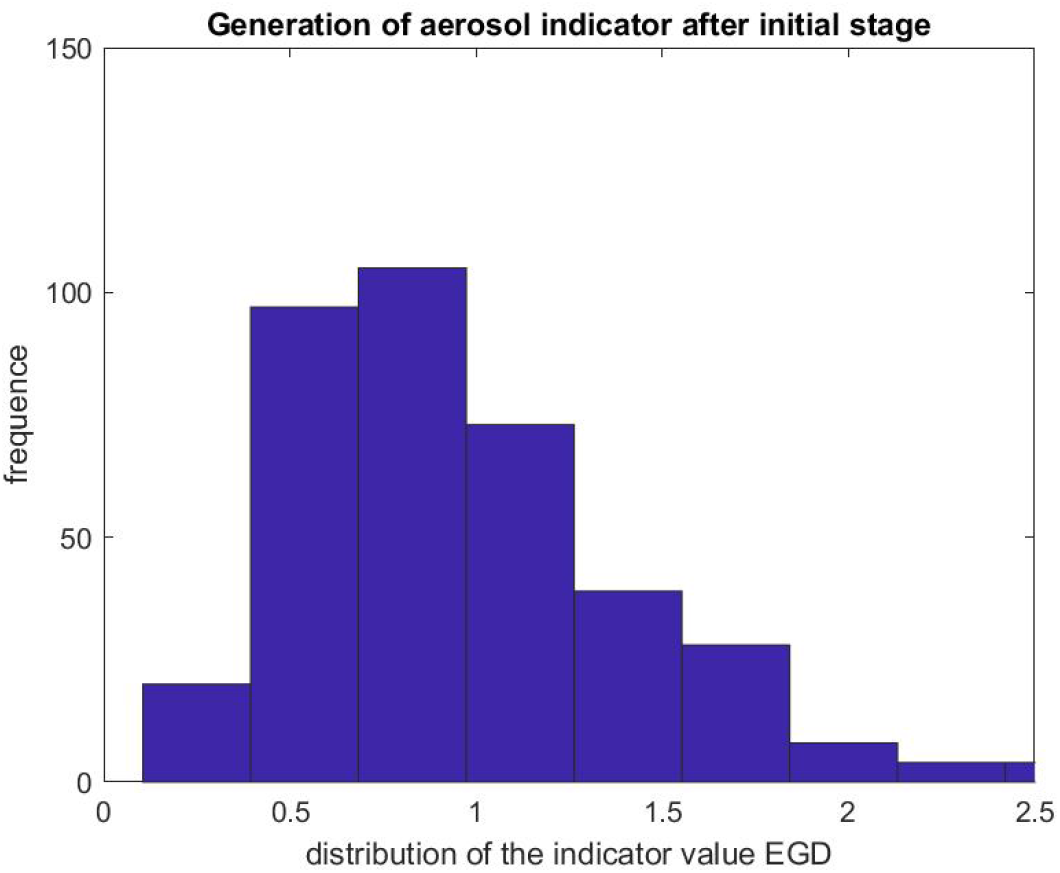
Indicator of aerosol generation for an EGD after the initial phase. The figure represent the distribution of the ratio 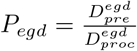

The risk of virus carried by small particles coming from the patient is therefore higher during the first few minutes of a colonoscopy and an EGD. However, this risk stays during the whole procedure for almost half of the procedures observed. This measurement supports the fact that PPE is essential for the staff.

Once the procedure is done and the surfaces are cleaned by the janitorial teams, one needs to make sure that most airborne particles generated by the patient procedures are evacuated by the HVAC system. We will discuss next the result of our clinical study.

### 3.2 Air Quality Turnover Time

As reported in the method section, we have systematically fitted an exponential decay function

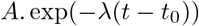

to the SPC curves in the time window of the turnover time between procedures, which is bounded by the highest peak value of the SPC (corresponding to the quick cleaning period of the room) and the lowest value that is below 50 *u_d_*, marking the *excellent* air quality level. *A* and *λ* are obtained with a least squares method. In principle, the procedural room door is closed during that time because the SPC in the hallway is much higher in our observation. Figure 11 shows the mean and standard variation of the *λ* value obtained from each fitting in each procedural room. We present the same calculation but for the large particle count using the same time interval. The *λ* value for small particle is smaller than the *λ* value for large particles which means that the relaxation time for small particles is longer than the relaxation time for large particle: as expected, the large particles are eliminated faster than small particles, which confirms the relative higher risk for SPC than Large Particle Count (LPC) and the importance of monitoring its value.

**Figure 11.**
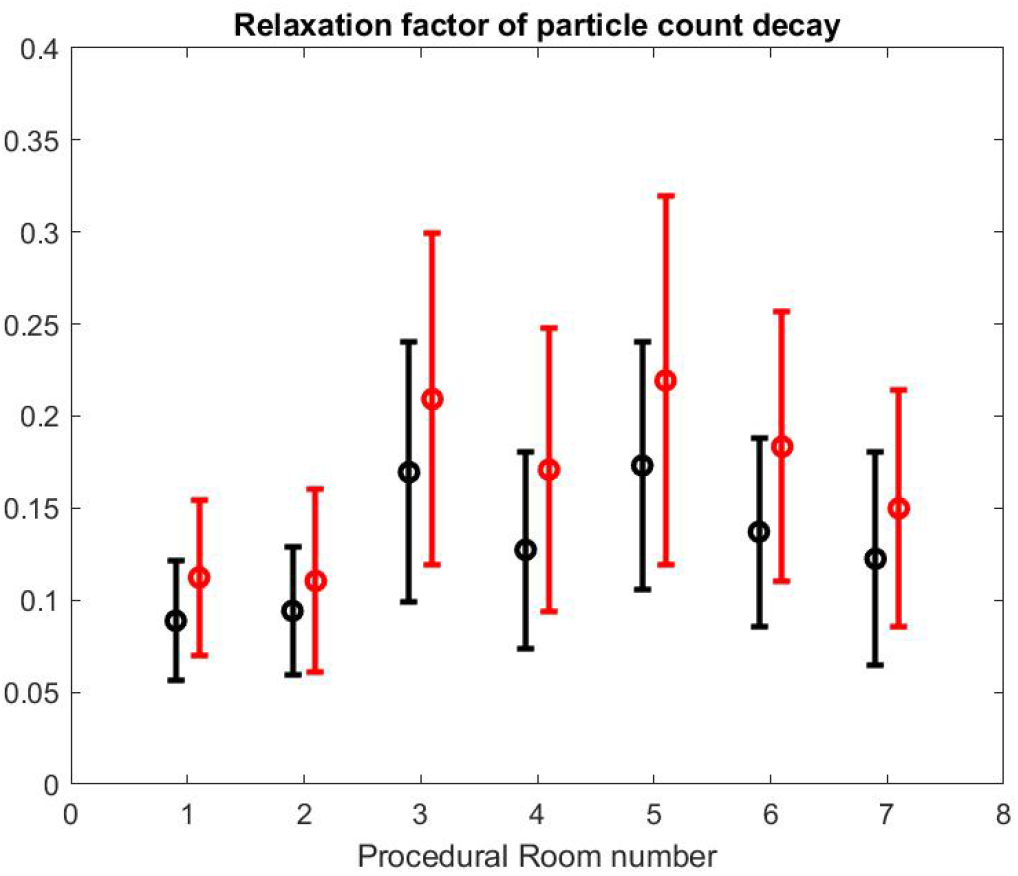
Relaxation factor *λ* of particle count decay *≈ exp*(*−λt*) for small particles (black) and large particles (red).

We can also notice that the decay factor depends on the procedural room itself. As a matter of fact, each procedural room may have a different structural size and inflow/outflow boundary conditions from the HVAC system.

Using the formula of [35] reported by the CDC guidelines for environmental infection control in health care facilities (2013) and measurement of inflow/outflow flux, one can obtain a theoretical prediction of the time it takes to the HVAC system to renew 99% of the air in the room. Figure 13 reports on this time estimate [33]. We can make a qualitative comparison between this theoretical estimate and the theoretical estimate based on a gross approximation of mass transfer of the air. We focus first on the relative comparison between each room air quality turnover to see if results of Figure 12 and Figure 13 are correlated. The relaxation factor we found for room 1 and 2 is about the same order and the smallest of all procedural rooms observed. Accordingly, room 1 and 2 in Figure 13 have the largest turnover time to renew the air. However, according to our measurements, room 7 renews air faster than the time given by the model’s formula. Room 4 and 5 seems to have a similar air quality turnover while in the theoretical estimate of Figure 13, room 5 has the best performances of all. Overall we observe a weak correlation between the theoretical prediction from [35] and our measurement of air quality turnover. We notice that the computed *λ* value varies significantly: according to computational fluid mechanics, the air quality renewal is dependent of the initial condition of air quality distributed in the 3-dimensional volume of the procedural room [9]. This initial condition may depend on a number of factors related to the dynamic of people leaving the procedural room, door motions and the amount of aerosol that has been generated.

**Figure 12.**
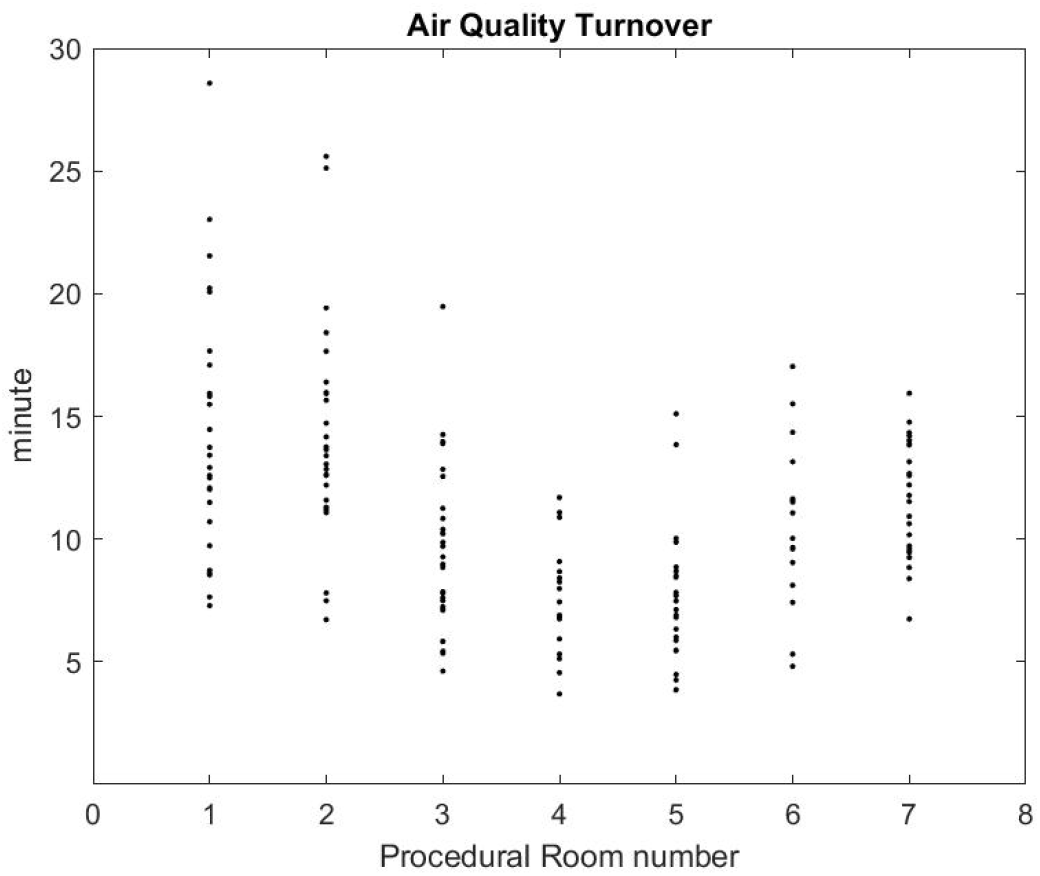
Air Quality Turnover Time measured in clinical conditions.

**Figure 13.**
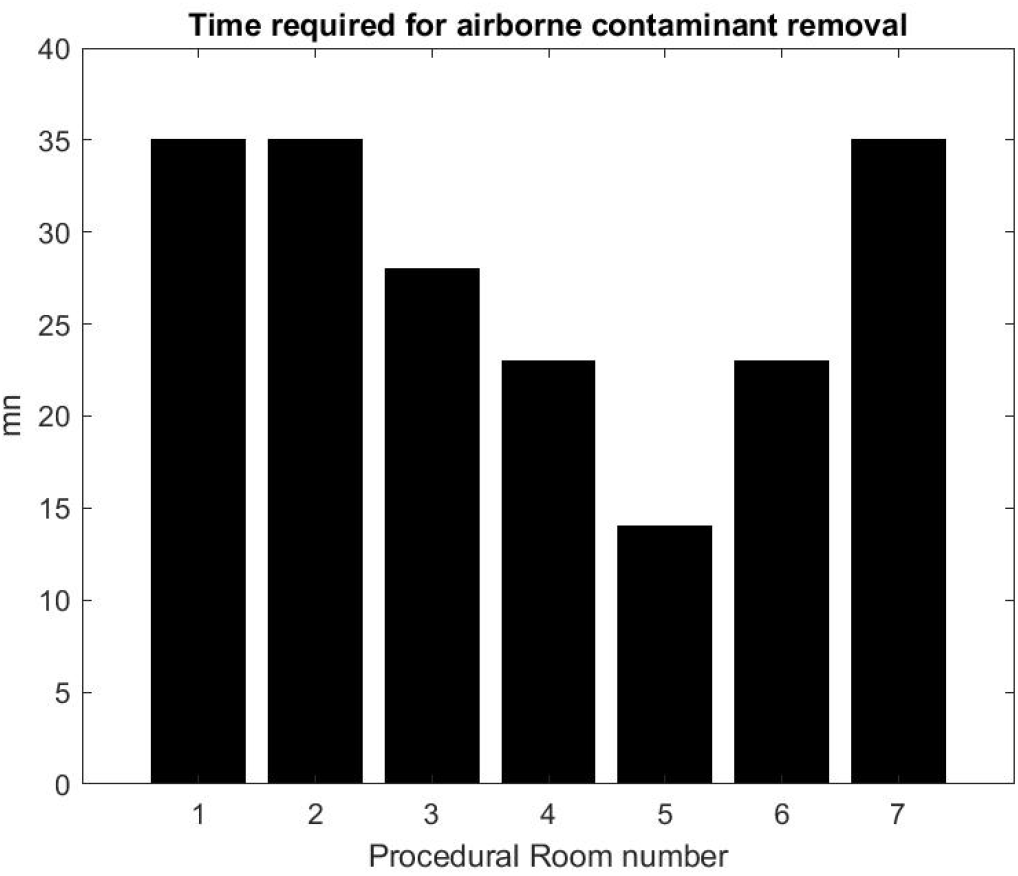
Turnover Time computed from the HVAC formula [35]

From the practical perspective, the most important factor is knowing how long it takes for the SPC to go back to an *excellent* air quality level that approximately corresponds to the baseline prior to any procedure. Figure 12 shows that on average, after the procedural room is closed it takes about 10 to 15 minutes to regain *excellent* air quality levels. While this is a much lower turnover time than the model’s prediction, the air quality turnover time associated with the measurement of SPC varies based on the room conditions, when the procedure is finished, and if the cleaning has been done [9]. The theoretical value provided by [35] can be very close to the worst case air-quality turnover observed inFigure 12.

On average, we estimate that reducing the turnover time from 30 minutes to 20 minutes based on air quality may increase the number of procedures during a shift from 7:00 am to 03:00 pm from 8 procedures to 10 procedures, which represents a 25% improvement on procedure throughput.

Let us next discuss the pros and cons of our method and results, and then conclude this paper.

## 4 Discussion and Conclusion

In our paper, we have attempted to provide a risk mitigation strategy for GI procedures in the context of COVID-19.

The main concept is that standard large volume procedures, such as colonoscopy or EGD, practiced in outpatient centers and gastroenterology departments, can expose staff and patients to an airborne virus. First and foremost, GI procedures have been classified as aerosol generating procedures [22,31]. COVID-19 has been found in patients’ stools as well as patients’ breath [34]. It has been shown in numerous studies that healthcare professionals are at a higher risk of infection than the general population [20]. While masks have their efficiency [21], long-term exposure to airborne contamination in closed environments increases the risk of contamination to the healthcare staff. In this context, we felt that it was important to assess the amount of small particles generated over a large number of GI procedures. We have presented a quantitative estimate based on a baseline consisting of the SPC prior to the procedures since we know that the presence of staff contributes to that measure through various mechanisms, breathing being one of the main sources. Our SPC ratio during/before the procedure constitutes a clear indicator of the level of aerosol generated by the procedure. Our findings also suggest that the initial phase of the procedure releases the most aerosol. Counting airborne particles does not prove or disprove the presence of an airborne virus; however, it is known that viruses can be carried around by a spray of such small particles that are constituted mostly of water and released by procedures generating aerosol [31]. Our study is the first systematic measurement in clinical conditions that confirms that both colonoscopy and EGD procedures generate significant level of aerosols.

Nevertheless, this result would be of limited value if it does not come along with a risk mitigation strategy that builds off of this information. The benefit of our cyber-physical infrastructure is that it continuously gives the SPC in every procedural room, giving clear information to healthcare staff that can help them minimize risk exposure. By introducing a concept of air quality turnover that follows each cleaning step of the room, our system helps to ensure that the door of the procedural room is properly closed as soon as possible in order to let the HVAC system renewing the air efficiently. As noticed earlier, any door opening or lack of sanitizing the room in time is easily detected by our system. We developed in our system a phone app to quickly and easily support the staff in their daily work. Over time and usage, it has become an autonomous, best-practice process that supports healthcare team work. We did not run into any technology acceptance of our system, due to the fact that our cyberinfrastructure does not add any work to the staff, nor require any type of training: the phone app is intuitive and gives only the information that matters when it matters.

Today, most clinics use a standard model formula based on room size and measured inflow/outflow HVAC to estimate what would be a reasonable amount of time needed to eliminate airborne particles. This is important because any healthcare professional should always ensure that the next patient going into a GI procedure will not be exposed to COVID-19 generated by any previous patient. As we know, asymptomatic patients might not be detected in time by today’s testing. As quoted by CDC, ”the times given assume perfect mixing of the air within the space (i.e., mixing factor = 1). However, perfect mixing usually does not occur. Removal times will be longer in rooms or areas with imperfect mixing or air stagnation”. To overcome these limitations, clinics use their own HVAC formulas with a very high level of elimination rate such as 99%. However, our results based on SPC shows that the air quality turnover widely varies from one procedure to another in the same room, and that HVAC efficiency in procedural rooms may not rank exactly as expected from the model formula, which is an over simplification of fluid mechanics and geometry. Their model formula can be miscalculated by: one, filling a room with numerous pieces of equipment that generate energy; two, the distribution of airborne particles at the moment the door is closed to let the HVAC renewal work may vary considerably; three, re-circulation zones and turbulent flows may completely change the results.

We think that our method based on new off-the-shelf sensor technology and real-time processing to communicate the information to healthcare staff is a more rigorous way to mitigate the risk from generated aerosols. Further, it turns out that our air quality turnover, while varying significantly from one case to another, is still far below the theoretical prediction based on the 99% elimination threshold given by HVAC system model formulas.

We conclude that our method may provide a safer and more efficient way of handling procedure turnover time in a clinic.

More work needs to be done; keeping record of contaminated cases to obtain long-term correlation between SPC measurement and potential transmission is certainly needed. We will argue, however, that a cyber-physical infrastructure like the one presented in this paper is a non-invasive and robust solution to eventually gain further validation by generating massive data and improving clinical practice.

## Data Availability

All data are available in the paper.

## Acknowledgment

We would like to thank Patrick Doolan for sharing his view with us on management and risk evaluation from his great experience acquired from the energy sector and Karen Schescke for her advise in GI clinical management.

